# Clinical modifiers of the association between type 1 diabetes and dementia incidence

**DOI:** 10.1101/2025.09.15.25335806

**Authors:** Anna M. Pederson, Peter Buto, Scott C Zimmerman, Kendra D. Sims, Audrey R. Murchland, Jingxuan Wang, M. Maria Glymour, Jennifer Weuve, Paola Gilsanz, Felicia Chi, Rachel A Whitmer, Alana T Brennan

**Author notes:** Funding: P01AG082653, F99AG083306, K99AG083121.

## Abstract

**Background:** Type 1 diabetes mellitus (T1DM) is associated with elevated dementia risk, but the mechanisms are not well understood. Prior studies suggest that co-occurring diabetes-related complications and other comorbidities may further increase dementia risk, but these studies are few and typically have small samples. Whether diabetes-related complications and other comorbidities modify the effect of T1DM on dementia risk remains unclear.

**Methods:** Data are from participants of the All of Us (AoU) cohort ages ≥ 50 years, with complete baseline surveys, linked electronic health records (EHRs), and either T1DM or no DM. Enrollment began in 2017, with data available through October 2023, including information prior to enrollment in AoU. Incident dementia was identified based on ICD-9, ICD-10, and SNOMED codes in participants’ EHRs. Baseline clinical comorbidities (diabetes complications and eye diseases, other vascular and metabolic comorbidities, and mental health conditions) were also identified using participants’ EHRs, classifying each as present if at least one diagnostic code occurred on or before the baseline survey.

**Results:** Among 232,429 participants (mean [SD] age 64.5 [9.0] years; 57.3% women), 2.3% had a T1DM diagnosis. Participants averaged 1.67 total comorbidities (SD = 2.08). T1DM and each comorbidity was associated with higher dementia incidence. T1DM was associated with higher dementia incidence among individuals with no comorbidities (HR = 1.77; 95% CI: 0.95-3.30), though the CI included 1. Each additional comorbidity increased risk (HR = 1.22; 95% CI: 1.19-1.26), with some evidence that the effect of T1DM differed by the number of comorbidities (HR = 0.94; 95% CI:0.86-1.01). The combined estimated effect of T1DM and most comorbidities was less than multiplicative. Depression was an exception; the dementia HR for individuals with both T1DM and depression (HR = 5.47; 95% CI: 4.23, 7.08) roughly reflected what would have been expected based on the HR for T1DM (HR = 2.01; 95% CI: 1.38, 2.92) times the HR for depression among those without T1DM (HR = 2.55; 95% CI: 2.22, 2.95).

**Conclusion:** Our findings suggest that T1DM and common comorbidities independently increase dementia risk, though their combined effects are generally less than multiplicative. However, depression in the context of T1DM is associated with major elevations in dementia risk.

## Introduction

The strong association between type 1 diabetes mellitus (T1DM) and dementia is not well understood, and in some studies appears greater than that observed for type 2 diabetes mellitus (T2DM).^1–3^ The limited available evidence suggests this risk may be amplified by co-occurring conditions, such as cardiovascular disease (CVD) and depression, reflecting the earlier onset of T1DM and longer duration of complications.^4–6^ Identifying such modifiers could clarify mechanisms and inform care for people with T1DM.

T1DM may influence dementia risk through multiple pathways, including diabetes related complications, cardiovascular and metabolic comorbidities, and behavioral or mental health. Established dementia risk factors such as hypertension, stroke, chronic kidney disease (CKD), neuropathy and hypoglycemia may contribute, though findings for retinopathy are mixed.^7,8–14^ Depression, common amongst individuals with T1DM may interact with glycemic control, creating a cycle of poorer self-care and further complications.^4^ These mechanisms may be particularly relevant given that complications often emerge earlier in life among those with T1DM.^15^

Despite these plausible pathways, existing studies are few and typically underpowered, with the largest including fewer than 4,000 individuals with T1DM.^4,8^ Using data from a large electronic health record (EHR) cohort, we evaluated whether comorbidities and complications modify the association between T1DM and incident dementia.

## Methods

### Study Sample

The All of Us (AoU) research program is a cohort of U.S. adults ≥ 18 years recruited through academic institutions and volunteer-based enrollment. At enrollment, participants completed surveys on sociodemographics, lifestyle, and health, and many consented to link past or future electronic health records (EHRs).^16,17^ Analyses were restricted to participants ≥ 50 years at baseline with linked EHRs. Follow-up through October 10, 2023, was available.

### Assessment of T1DM

We used a validated algorithm to classify participants as having T1DM if they had ≥1 EHR encounter with a T1DM diagnostic code.^3^ Those without diabetes codes were classified as not having diabetes; individuals with T2DM but not T1DM were excluded.

### Modifiers

Baseline comorbidities and complications were identified from EHRs, including glaucoma, cataracts, retinopathy, neuropathy, hyper/hypoglycemic events, congestive heart failure (CHF), stroke, CKD, myocardial infarction (MI), hypertension, dyslipidemia, obesity, alcohol use disorder (AUD), anxiety, and depression. Participants were classified as having a condition if ≥1 diagnostic code was present before the baseline survey. Obesity was also assessed using measured BMI, with WHO criteria (≥30 kg/m^2^).^18^

We created four composite measures reflecting potential pathways linking comorbidities to dementia risk: (1) diabetes complications and eye diseases, including glaucoma, cataracts, retinopathy, neuropathy, and hyper/hypoglycemic events (range 0–6); (2) cardiovascular and metabolic comorbidities, including dyslipidemia, hypertension, stroke, MI, CHF, obesity, and CKD (range 0–7); (3) mental health conditions, including AUD, anxiety, and depression (range 0–3); and (4) overall comorbidity burden, defined as the total number of conditions across groups 1–3 (range 0–15). Nearly all people with T1DM who developed dementia had hypertension or dyslipidemia at enrollment. Following the All of Us reporting guidelines, which restricts reporting samples of less than 20 participants, we did not estimate separate models for hypertension and dyslipidemia. Both were included in the summary measures.

### Assessment of Dementia

Dementia was defined as Alzheimer’s disease (AD), vascular dementia, or dementia of unknown etiology, consistent with our prior work.^3^ Incident cases were identified from the first ICD-9, ICD-10, or SNOMED code after baseline. Participants with a prior dementia diagnosis were excluded, and follow-up was censored at death or the last EHR encounter recorded in the data (October 1, 2023).

### Covariates

We estimated the association between T1DM and incident dementia using models that adjusted for demographic covariates reflecting potential confounding, as described previously.^3^ All covariates were self-reported at baseline and included age, sex/gender (male or female), race and ethnicity (Non-Hispanic White, Hispanic/Latino, or Other), educational attainment (less than high school, high school, some college, or college and above), and household income (adjusted by dividing by the square root of household size).

### Statistical Analysis

We summarized baseline demographic and clinical characteristics of participants with and without T1DM. We used Cox proportional hazards models, with time from baseline as the time scale, to estimate hazard ratios (HRs) for the association of T1DM versus no diabetes with dementia. For each comorbidity, we fit models including T1DM, the comorbidity, and their interaction term to test effect modification. All models were adjusted for age, sex/gender, race and ethnicity, education, and household income.

Effect modification was assessed on a multiplicative scale, where the interaction term evaluates whether the combined effect of T1DM and the comorbidity is greater than (interaction HR ≤1), equal to (interaction HR =1), or less than (interaction HR <1) the product of their individual effects. We assessed two distinct questions: the effect of each modifier among people with T1DM; and whether the effect of the co-occurrence of T1DM and each modifier was greater or less than what would be expected based on their individual hazard ratios (i.e., comparing the interaction term to 1).

To estimate the effect of each modifier among people with T1DM, combined HRs and 95% CIs were calculated as the sum of log-HRs for each comorbidity and its interaction with T1DM, using methods described by Wang and Husnik.^19^

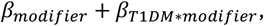

where *β*_*modifier*_ is the coefficient for the comorbidity and *β*_*T1DM*\**modifier*_is the coefficient for its interaction with T1DM. The variance of this sum was derived from the model output’s variance-covariance matrix:

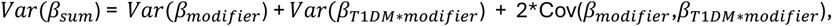

and the 95% CI was calculated by exponentiating the sum of the coefficients +/-1.96 times the square root of the sum of the variance:

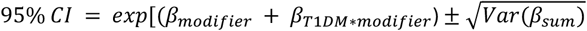

## Results

In our sample of 232,429 participants, 2.3% were classified as having T1DM. Over a mean of 2.25 years of follow-up, 1,416 (0.61%) participants developed dementia. The mean age was 64.5 years (SD 9.0), 57.3% were female, and 50.0% had at least a college degree (Table 1). Participants had, on average, 1.67 total comorbidities (SD 2.08). This included a mean of 0.45 (SD 0.84) diabetes complications or eye conditions, 0.88 (SD 1.12) other CVD and metabolic comorbidities, and 0.34 (SD 0.70) mental health conditions.

**Table 1.** Characteristics of the analytic sample, by DM status.

|  | All (n=232429) | Type 1 DM<br>(n=5444) | No Diabetes<br>(n=226985) |
| --- | --- | --- | --- |
| **Age (years), mean (SD)** | 64.54 (8.98) | 65.11 (8.72) | 64.53 (8.99) |
| **Female, n (%)** | 133143 (57.3%) | 2742 (50.4%) | 130401 (57.4%) |
| **Race and ethnicity** |  |  |  |
| White | 145982 (62.8%) | 2598 (47.7%) | 143384 (63.2%) |
| Latino | 28293 (12.2%) | 1030 (18.9%) | 27263 (12%) |
| Other | 58154 (25%) | 1816 (33.4%) | 56338 (24.8%) |
| **Educational attainment** |  |  |  |
| Advanced Degree | 61645 (26.5%) | 826 (15.2%) | 60819 (26.8%) |
| College Graduate | 54714 (23.5%) | 1059 (19.5%) | 53655 (23.6%) |
| Some College | 57039 (24.5%) | 1590 (29.2%) | 55449 (24.4%) |
| High School or GED | 35749 (15.4%) | 1074 (19.7%) | 34675 (15.3%) |
| Less than High School | 17406 (7.5%) | 739 (13.6%) | 16667 (7.3%) |
| **BMI (kg/m^2)** | 29.38 (6.94) | 32.05 (7.43) | 29.3 (6.91) |
| **Income (divided by the square root of household size)** | 60371.17<br>(50977.87) | 40399.9<br>(41427.05) | 60815.82<br>(51081.54) |
| **Ever smoked (100 Cigarettes lifetime)** | 98869 (42.5%) | 2474 (45.4%) | 96395 (42.5%) |
| **Glaucoma** | 12839 (5.5%) | 1376 (25.3%) | 11463 (5.1%) |
| **Retinopathy** | 2961 (1.3%) | 1325 (24.3%) | 1636 (0.7%) |
| **Cataracts** | 27833 (12%) | 2731 (50.2%) | 25102 (11.1%) |
| **Neuropathy** | 49207 (21.2%) | 4094 (75.2%) | 45113 (19.9%) |
| **Heart failure** | 6415 (2.8%) | 1379 (25.3%) | 5036 (2.2%) |
| **Stroke** | 6984 (3%) | 937 (17.2%) | 6047 (2.7%) |
| **Myocardial Infarction** | 6548 (2.8%) | 1080 (19.8%) | 5468 (2.4%) |
| **Chronic kidney disease** | 11477 (4.9%) | 2256 (41.4%) | 9221 (4.1%) |
| **Obesity** | 45983 (19.8%) | 1994 (36.6%) | 43989 (19.4%) |
| **Hyperglycemic event** | 9533 (4.1%) | 3085 (56.7%) | 6448 (2.8%) |
| **Hypoglycemic event** | 2291 (1%) | 895 (16.4%) | 1396 (0.6%) |
| **Alcohol Use Disorder** | 9039 (3.9%) | 664 (12.2%) | 8375 (3.7%) |
| **Anxiety** | 38303 (16.5%) | 2866 (52.6%) | 35437 (15.6%) |
| **Depression** | 31368 (13.5%) | 2039 (37.5%) | 29329 (12.9%) |
| **Diabetes Complications and Eye Diseases<sup>a</sup>** | 0.45 (0.84) | 2.48 (1.41) | 0.40 (0.75) |
| **Other Vascular and Metabolic Comorbidites<sup>b</sup>** | 0.88 (1.12) | 3.13 (1.50) | 0.83 (1.05) |
| **Mental Health Conditions<sup>c</sup>** | 0.34 (0.70) | 1.02 (0.97) | 0.32 (0.69) |
| **Sum of Overall Comorbidities<sup>d</sup>** | 1.67 (2.08) | 6.63 (2.76) | 1.55 (1.91) |

[a] Diabetes complications and eye diseases: Total number of diabetes-related complications including glaucoma, cataracts, retinopathy, neuropathy, and hyper- and hypo- glycemic events. [b] Other vascular and metabolic comorbidities: Total number of vascular and metabolic comorbidities, including dyslipidemia, hypertension, stroke, myocardial infarction, congestive heart failure, obesity, and chronic kidney disease. [c] Mental health conditions: Count of alcohol use disorder, anxiety, and depression. [d] Sum of overall comorbidities: Total number of comorbidities present at baseline across all comorbidities listed in groups [a], [b], and [c].

Table 2 shows HRs and 95% CIs for dementia risk by comorbidity among individuals with T1DM, comparing those with versus without each condition. Within the T1DM group, chronic kidney disease (HR 1.77; 95% CI: 1.18–2.65), anxiety (HR 1.70; 95% CI: 1.14–2.53), and depression (HR 2.72; 95% CI: 1.76–4.20) were significantly associated with higher dementia incidence. Individuals with co-occurring T1DM and each of these comorbidities were at markedly elevated risk (e.g., HR for co-occurring T1DM and depression HR = 5.47; 95% CI: 4.23, 7.08) compared to individuals with neither. Among people with T1DM, the estimated effect of each comorbidity ranged from HR 1.10 (glaucoma) to HR 2.72 (depression), calculated as the sum of the comorbidity main effect and its interaction with T1DM; however, for most comorbidities the confidence interval included 1 (see Methods, Table 2).

**Table 2.** Dementia Hazard Ratios for each Modifier Among Individuals with Type 1 Diabetes Mellitus.

| Modifier | Dementia cases / among people without the modifier | Dementia cases / among people with the modifier | Dementia hazard ratio (95% confidence interval) |
| --- | --- | --- | --- |
| **Glaucoma** (ref: no glaucoma) | 103/4067 | 41/1375 | 1.10 (0.72, 1.68) |
| **Cataracts** (ref: no cataracts) | 49/2712 | 95/2730 | 1.22 (0.81, 1.86) |
| **Retinopathy** (ref: no retinopathy) | 101/4117 | 43/1325 | 1.31 (0.86, 2.00) |
| **Neuropathy** (ref: no neuropathy) | 31/1350 | 113/4092 | 1.17 (0.71, 1.91) |
| **Hyperglycemic Event** (ref: no hyperglycemic event) | 59/2357 | 85/3085 | 1.45 (0.97, 2.17) |
| **Hypoglycemic Event** (ref: no hypoglycemic event) | 113/4547 | 31/895 | 1.34 (0.82, 2.19) |
| **Congestive Heart Failure** (ref: no congestive heart failure) | 96/4063 | 48/1379 | 1.13 (0.74, 1.72) |
| **Stroke** (ref: no stroke) | 98/4505 | 46/937 | 1.46 (0.94, 2.26) |
| **Myocardial Infarction** (ref: no myocardial infarction) | 106/4362 | 38/1080 | 1.14 (0.72, 1.80) |
| **Chronic Kidney Disease** (ref: no chronic kidney disease) | 66/3187 | 78/2333 | 1.77 (1.18, 2.65) |
| **Obesity** (ref: no obesity) | 89/3448 | 55/1994 | 1.01 (0.67, 1.52) |
| **Alcohol Use Disorder** (ref: no alcohol use disorder) | 122/4778 | 22/664 | 1.37 (0.78, 2.43) |
| **Anxiety** (ref: no anxiety) | 85/3405 | 59/2037 | 1.70 (1.14, 2.53) |
| **Depression** (ref: no depression) | 47/2577 | 97/2865 | 2.72 (1.76, 4.20) |
| **Diabetes Complications and Eye Diseases<sup>a</sup>, per complication** |  |  | 1.16 (1.00, 1.34) |
| **Other Vascular and Metabolic Comorbidities<sup>b</sup>, per comorbidity** |  |  | 1.17 (1.03, 1.34) |
| **Mental Health Conditions<sup>c</sup>, per condition** |  |  | 1.53 (1.25, 1.88) |
| **Sum of Overall Comorbidities<sup>d</sup>, per comorbidity** |  |  | 1.15 (1.07, 1.24) |

Although point estimates for both T1DM and each comorbidity were associated with elevated hazard of dementia (i.e., HR≤1), the combined effects of T1DM and most comorbidities were less than multiplicative. For example, the T1DM-CHF interaction was <1.0, suggesting that the estimated effect of T1DM was smaller among individuals with CHF (HR 1.60; 95% CI: 1.06, 2.41) compared to the estimated effect of T1DM among those without CHF (HR 2.82; 95% CI: 2.18, 3.63) (Figure 1). Interactions were largely below 1 with wide CIs, except relative effects of T1DM appear smaller among those with MI (HR 0.56; 95% CI: 0.35, 0.99), stroke (HR 0.62; 95% CI: 0.38, 1.00), and hypoglycemic events (HR 0.36; 95% CI: 0.19, 0.68) (Supplemental Table 1).

**Figure 1.**
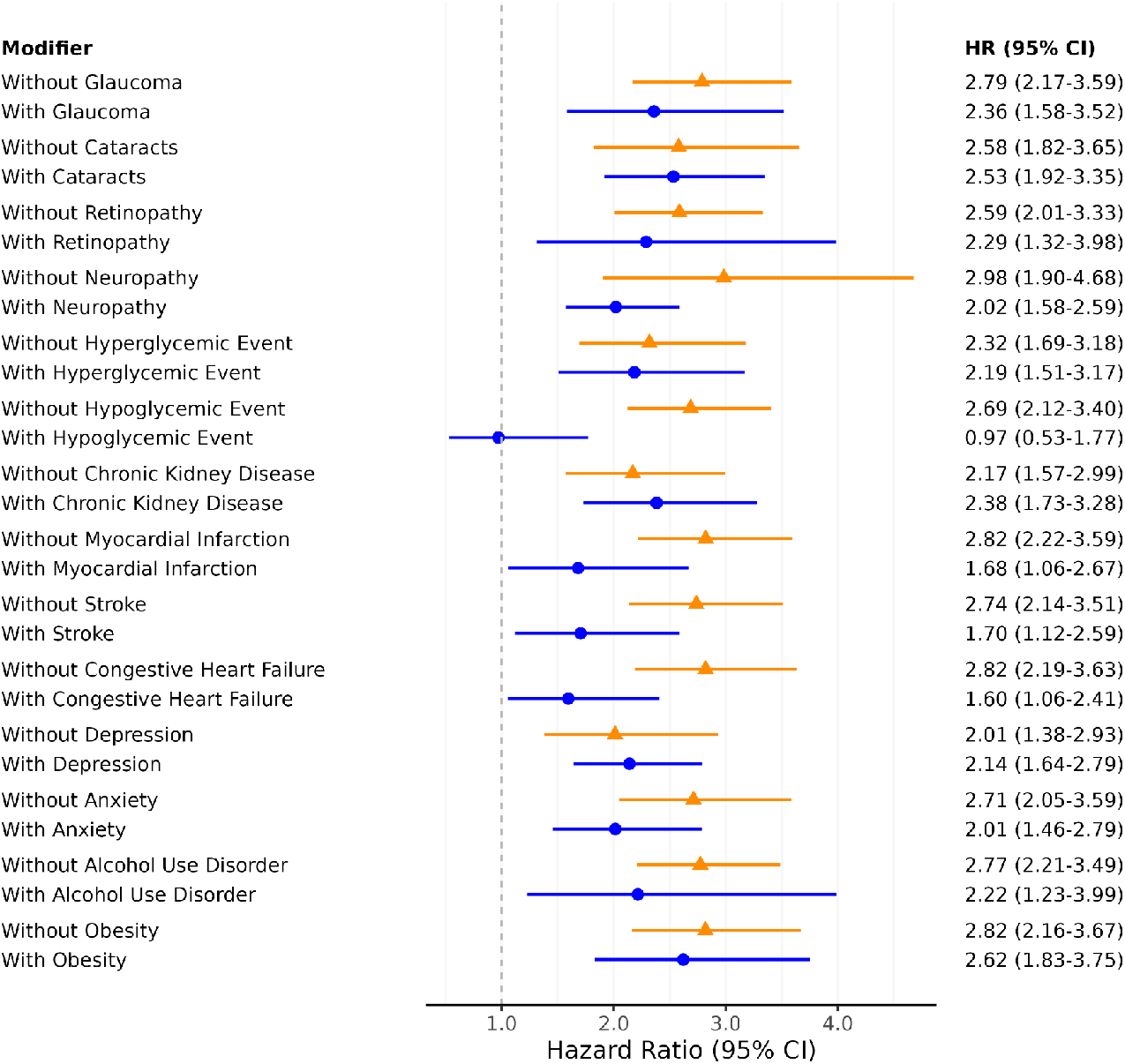
Association of Type 1 Diabetes with incident Dementia, among individuals with (blue) or without (orange) each modifier. Associations and 95% confidence intervals for type 1 diabetes (T1DM) with dementia risk by modifier status. Estimates in orange show the main effect of T1DM in individuals without the modifier. Estimates in blue show the main effect of T1DM in individuals with the modifier. All models were adjusted for age, race and ethnicity, gender, education, and income.

In addition, comorbidity burden was positively associated with dementia incidence. For example, among people without T1DM, each additional comorbidity in the sum of the overall count was associated with a 22% increase in dementia incidence (HR 1.22; 95% CI: 1.19-1.26). Similar patterns were observed for each additional vascular and metabolic comorbidity (HR 1.36; 95% CI: 1.30-1.43), diabetes complications or eye condition (HR 1.27; 95% CI: 1.19-1.35), and mental health comorbidity (HR 1.66; 95% CI: 1.54-1.79) (Supplemental Table 1).

## Discussion

To our knowledge, this is among the first large EHR-based studies to examine a broad set of clinical and behavioral comorbidities as potential modifiers of the association between T1DM and incident dementia. We found that both T1DM and nearly all comorbidities were independently associated with elevated dementia risk, highlighting that each condition on its own contributes meaningfully to risk. However, most interaction estimates in our study were <1, indicating that the combined effect was smaller than anticipated based on multiplying the relative effects of T1DM and the comorbidity. The estimated effect of co-occurring T1DM and depression was especially high, indicating over a 5-fold excess. Many confidence intervals for interactions included 1, providing no clear evidence that the co-occurrence of T1DM and a comorbidity was associated with dementia risk beyond what would be expected from their individual effects.

Although interpretations about mechanisms are speculative, sub-multiplicative interactions (where the combined effect of two risk factors is smaller than what would be expected if their effects multiplied together) may indicate that effects are operating through similar biological pathways and either risk factor is sufficient to complete that step of the mechanism. In contrast, interaction terms near 1, as observed for cataracts, hyperglycemic events, CKD, and depression, may suggest independent mechanisms contributing to dementia risk.

Our study has important strengths. We used a validated EHR-based algorithm to accurately identify individuals with T1DM^3^, enabling a large sample, and leveraged EHR data to capture comorbidities that may be underreported in self-reported surveys. We also examined a wide range of clinical and behavioral comorbidities, providing a comprehensive assessment of factors influencing dementia risk in T1DM. Limitations should also be noted. T1DM and comorbidities may be misclassified in EHRs.^3^ Comorbidities may be underreported if care occurred outside contributing systems or if conditions developed after baseline, potentially underestimating associations with dementia risk. Comorbidities may be differentially captured by diabetes status, as individuals with T1DM are more likely to engage with health care and receive diagnoses; this is unlikely to bias analyses restricted to T1DM but may influence interaction estimates. We did not attempt to evaluate mediation, though every comorbidity we evaluated was far more common among people with T1DM than people without. Finally, temporality is uncertain, as our definition of comorbidity did not distinguish long-standing conditions from new diagnoses that may reflect prodromal dementia. A critical question for future research is how treatment or management modify the T1DM–dementia association.

Overall, our results suggest that nearly every comorbid condition was associated with some increase in dementia risk among people with T1DM, with particularly large excesses associated with chronic kidney disease, anxiety, and depression. These increases were generally not more than what would be expected if T1DM and the comorbid condition independently influence dementia risk.

## Data Availability

The data used in this study are available through the All of Us Researcher Workbench (https://www.researchallofus.org/data-tools/workbench/), which requires institutional approval and researcher compliance with data use policies. Due to data use agreements and participant privacy, individual-level data cannot be shared publicly. Code used for the analysis is available upon request.

https://www.researchallofus.org

