## Supplementary Material for "Clinical modifiers of the association between type 1 diabetes and dementia incidence"

**eTable 1.** Dementia Hazard Ratios for Type 1 Diabetes, Modifiers, and Their Interaction

**eTable 2.** Supplemental Table 2. Concept IDs* Used for Each Comorbidity

**Supplemental Table 1. Dementia Hazard Ratios for Type 1 Diabetes, Modifiers, and Their Interaction**

| | Dementia hazard ratio (95% confidence interval) associated with type 1 diabetes, comorbidity, and the interaction (referent is individuals with neither diabetes nor comorbidity) |
|---|---|
| Comorbidity | |
| **Glaucoma** | |
| Type 1 Diabetes | 2.78 (2.16, 3.58) |
| Glaucoma | 1.30 (1.07, 1.58) |
| Glaucoma:T1DM | 0.84 (0.53, 1.35) |
| **Cataracts** | |
| Type 1 Diabetes | 2.58 (1.82, 3.65) |
| Cataracts | 1.25 (1.07, 1.45) |
| Cataracts:T1DM | 0.98 (0.63, 1.52) |
| **Retinopathy** | |
| Type 1 Diabetes | 2.58 (2.00, 3.33) |
| Retinopathy | 1.48 (0.96, 2.29) |
| Retinopathy:T1DM | 0.88 (0.48, 1.62) |
| **Neuropathy** | |
| Type 1 Diabetes | 2.98 (1.90, 4.67) |
| Neuropathy | 1.72 (1.51, 1.98) |
| Neuropathy:T1DM | 0.67 (0.40, 1.12) |
| **Hyperglycemic Event** | |

| | |
|---|---|
| Type 1 Diabetes | 2.39 (1.69, 3.17) |
| Hyperglycemic Event | 1.54 (1.18, 2.03) |
| Hyperglycemic Event:T1DM | 0.94 (0.58, 1.52) |
| **Hypoglycemic Event** | |
| Type 1 Diabetes | 2.68 (2.12, 3.40) |
| Hypoglycemic Event | 3.71 (2.45, 5.62) |
| Hypoglycemic Event:T1DM | 0.36 (0.19, 0.68) |
| **Congestive Heart Failure (CHF)** | |
| Type 1 Diabetes | 2.82 (2.18, 3.63) |
| CHF | 2.00 (1.59, 2.51) |
| CHF:T1DM | 0.56 (0.35, 0.91) |
| **Stroke** | |
| Type 1 Diabetes | 2.73 (2.13, 3.50) |
| Stroke | 2.34 (1.91, 2.87) |
| Stroke:T1DM | 0.62 (0.38, 1.00) |
| **Myocardial Infarction (MI)** | |
| Type 1 Diabetes | 2.82 (2.21, 3.59) |
| MI | 1.91 (1.51, 1.98) |
| MI:T1DM | 0.56 (0.35, 0.99) |
| **Chronic Kidney Disease (CKD)** | |
| Type 1 Diabetes | 2.17 (1.57, 2.99) |
| CKD | 1.61 (1.32, 1.97) |

| | |
|---|---|
| CKD:T1DM | 1.09 (0.70, 1.71) |
| **Obesity** | |
| Type 1 Diabetes | 2.81 (2.16, 3.67) |
| Obesity | 1.09 (0.91, 1.30) |
| Obesity:T1DM | 0.93 (0.59, 1.44) |
| **Alcohol Use Disorder (AUD)** | |
| Type 1 Diabetes | 2.77 (2.20, 3.48) |
| AUD | 1.72 (1.30, 2.28) |
| AUD:T1DM | 0.79 (0.42, 1.49) |
| **Anxiety** | |
| Type 1 Diabetes | 2.71 (2.05, 3.58) |
| Anxiety | 2.29 (1.97, 2.67) |
| Anxiety:T1DM | 0.74 (0.48, 1.13) |
| **Depression** | |
| Type 1 Diabetes | 2.01 (1.38, 2.92) |
| Depression | 2.55 (2.22, 2.95) |
| Depression:T1DM | 1.06 (0.67, 1.67) |
| **Diabetes Complications and Eye Diseases**[a] | |
| Type 1 Diabetes | 2.23 (1.40, 3.55) |
| Diabetes Complications and Eye Diseases | 1.27 (1.19, 1.35) |
| Diabetes Complications and Eye Disease:T1DM | 0.91 (0.78, 1.06) |
| **Other Vascular and Metabolic Comorbidities**[b] | |

| | |
|---|---|
| Type 1 Diabetes | 2.56 (1.50, 4.38) |
| Vascular Comorbidities | 1.36 (1.30, 1.43) |
| Vascular Comorbidities:T1DM | 0.86 (0.75, 0.99) |
| **Mental Health Conditions**[c] | |
| Type 1 Diabetes | 2.32 (1.66, 3.26) |
| Mental Health Comorbidities | 1.66 (1.54, 1.79) |
| Mental Health Comorbidities:T1DM | 0.92 (0.74, 1.14) |
| **Sum of Overall Comorbidities**[b] | |
| Type 1 Diabetes | 1.77 (0.95, 3.30) |
| Sum of Comorbidities | 1.22 (1.19, 1.26) |
| Sum of Comorbidities:T1DM | 0.94 (0.86, 1.01) |

All models are adjusted for age, gender, race and ethnicity, educational attainment, and household income. [a] Diabetes complications and Eye Diseases: Total number of diabetes-related complications including glaucoma, cataracts, retinopathy, neuropathy, and hyper- and hypo- glycemic events. [b] Other vascular and metabolic comorbidities: Total number of vascular and metabolic comorbidities, including dyslipidemia, hypertension, stroke, myocardial infarction, congestive heart failure, obesity, and chronic kidney disease. [c] Mental health conditions: Count of alcohol use disorder, anxiety, and depression. [d] Sum of overall comorbidities: Total number of comorbidities present at baseline across all comorbidities listed in groups [a], [b], and [c].

**Supplemental Table 2. Concept IDs* Used for Each Comorbidity**

| Comorbidity | Concept ID(s) | Notes |
|---|---|---|
| Glaucoma | 437541, 44826578, 1568761, 441561, 44828911, 156769, 441284, 44834658, 1568768, 435543, 44819624, 1568792, 425262 | Includes descendants and all mapped codes |
| Cataracts | 1568646, 44830011, 375545, 44828918, 44830014, 44837042, 44828917, 35207548, 44823055, 44824186, 44830013, 44837041, 1568667, 44828913, 44833484, 1568670, 44831167, 44827724, 44830010, 44826580, 44828915, 44837043, 35207544, 44834663, 44831166, 44820797, 44835867, 44831165, 44819629, 44828914, 44833483, 44819631, 44828916, 1568664, 44830012, 44827725, 35207545 | |
| Retinopathy | 44831148, 44833466, 44827714, 45538261, 44820785, 44833465, 1568707, 44835858 | |
| Neuropathy | 44827615, 44825331, 1568864, 35207451, 44822493, 44822492, 44826006, 44827706, 44828884, 35207477, 44829067, 4301699, 44827690, 44825349, 1568865, 44826563 | |
| Hyperglycemic Event | 4214376 | |
| Hypoglycemic Event | 24609, 44831048, 35206889, 380688, 35206888, 44820686 | |
| Congestive Heart Failure | 4023479, 44782655, 4242669, 4229440, 319835, 314378 | |
| Stroke | 1569193, 1569190, 1569184, 35207403, 35207404, 35207405, 35207406, 35207407, 44820872, | |

| | |
|---|---|
| | 44835946, 44820873, 44824253, 443454, 432923 |
| Myocardial Infarction | 4329847, 312327, 1326591, 1569126 |
| Chronic Kidney Disease | 46271022, 44830172, 1571486, 443614, 443601, 443597, 443612, 443611, 35209274, 35209275, 35209276, 725440, 725441, 725442, 35209277, 35209278, 44837191, 44830173, 44827888, 44820970, 44837192 |
| Alcohol Use Disorder | 44824125, 44826511, 433753, 44820730, 1568096, 45581404, 45600692, 45571707, 45538006, 1568097, 45576493, 45557154, 45576494, 1568099, 45576495, 725273, 725274, 725275, 44826518, 44829936, 45595845, 44828841, 725271, 435243, 1568100, 44832244, 45566777, 45562002, 45595847, 45566776, 45600695, 45562003, 45557155, 45571708, 45600696, 45591082, 45586189, 45586188, 45538007, 45571709, 45591084, 45605452, 45552431, 45566775, 45586190, 45533065, 45586187, 1568102, 45566781, 45542791, 45566780, 45547675, 1568104, 45591087, 45586192, 45562005, 1568103, 45566778, 45595851, 45552432, 45576497, 1568105, 45605456, 725276, 725278, 725277, 725280, 37402462, 37402461, 44826487, 44835774, 44824107, 44824108, 44832220, |

| | |
|---|---|
| | 44836956, 44819550, 44825305, 44819551, 44831100, 44831080 |
| Anxiety | 442077, 440374, 4304010, 4152371 |
| Depression | 438727, 440696, 766349, 440383, 4282096, 4195572, 4049623, 444100, 4098302, 4282316, 4141454, 433991, 432883, 434911, 438406, 441534, 435220 |

* Concept IDs are defined based on SNOMED codes and ICD-9 or ICD-10 codes and are used in All of Us to identify clinical conditions from electronic health records.
